# Prevalence and risk factors of elevated ALT in 3,399 treatment-naïve HBV/HDV co-infected patients: A comparison with propensity score-matched HBV mono-infected patients

**DOI:** 10.1101/2025.04.11.25325563

**Authors:** Habiba Kamal, Ganbolor Jargalsaikhan, Sanjaasuren Enkhtaivan, Karin Lindahl, Hannes Hagström, Daniel Bruce, Michael Ingre, Bekhbold Dashtserenx, Oyungerel Lkhagva-Ochir, Tuvshinjargal Ulziibadrakh, Andreas Bungert, Heiner Wedemeyer, Naranjargal B Dashdorj, Soo Aleman

## Abstract

**Background:** Chronic hepatitis D (CHD) causes severe chronic hepatitis. Knowledge is limited about factors correlating with ALT in treatment-naïve patients with CHD. This study analyzed the prevalence and determinants of ALT elevation in a large cohort of patients with CHD, including young adults, compared to propensity score-matched (PSM) HBV mono-infected patients.

**Methods:** We identified 3,399 treatment-naïve HBsAg+ adults with CHD (HDV RNA positive) or 2,556 with HBV mono-infection attending a liver center in Mongolia during 2015-2023. The relation between ALT levels, virological, biochemical and fibrosis parameters were assessed using Spearman correlation coefficient (rho). Logistic regression analysis was used to identify factors associated with elevated ALT in the whole cohort, and in 2,231 PSM on age, sex, and date of initial test pairs with CHD and mono-HBV.

**Results:** In CHD, 78.5% of patients had ALT elevation, with the highest prevalence in the 18-29 years group (n=340, 84%). This age group displayed 4.80-odds ratio (OR) for elevated ALT, 2.76-OR for elevated GGT, and 5.08-OR for cirrhosis, than matched mono-HBV group (all p<0.05). ALT levels correlated weakly with HDV RNA (rho=0.23) and liver stiffness (rho=0.37), moderately with GGT (rho=0.48), while showed no correlation with HBV DNA or HBsAg. Independent factors for elevated ALT were age <30 years, elevated GGT and HDV RNA ≥100,000 IU/mL.

**Conclusions:** In this large cohort of Asian patients with CHD, an early and more severe inflammatory process regardless of liver cirrhosis could be demonstrated in CHD compared to HBV-monoinfection supporting early administration of anti-HDV therapy.

## Introduction

Chronic hepatitis D (CHD) is associated with an aggressive liver disease course [1]. HBV/HDV co-infection affects nearly 9 to 19 million people worldwide [2]. The prevalence is highest in Mongolia, Amazon basin and West Africa, while generally low (<0.2%) in Western countries, with the majority consisting of migrants from endemic regions [2,3]. CHD is classified as a rare disease in the European Union, with orphan designation when developing new HDV drugs [4]. At diagnosis, 30-50% of patients with CHD present with advanced liver disease, at a mean age of 40 years [5,6].

Clinical evaluation of HBV/HDV co-infected patients involves biochemical markers such as alanine aminotransferase (ALT), virological parameters, and fibrosis assessment to guide disease staging, prognosis and inform treatment decisions [7]. Elevated ALT level reflects hepatic inflammatory process, and correlates with liver-related outcomes such as cirrhosis and hepatocellular carcinoma (HCC) [8]. Hence, ALT normalization is an accepted surrogate marker for improved clinical outcomes in HDV drug trials, but some discrepancies between ALT and HDV RNA responses have been observed with recently introduced bulevirtide therapy [9,10]. Knowledge about ALT levels is therefore crucial to understand the severe pathological process and for the management of CHD, particularly in the light of new HDV drugs in development.

While ALT elevation is frequent in CHD, data on its prevalence, severity, and association with patients characteristics and other factors across the natural course of CHD in untreated patients remain scarce [6,11,12]. Particularly data on young adults with CHD compared to chronic hepatitis B (CHB) is absent. Studies on CHD have been constrained by small sample size, inclusion of both treatment-naïve and experienced patients, and tertiary care settings, reducing statistical power and introducing potential bias [6,11,12].

To address this knowledge gap, we analyzed the prevalence and independent factors for ALT elevation in HBV/HDV co-infected patients, including young adults, and compared them to propensity score-matched (PSM) HBV mono-infected patients. This analysis was enabled using a unique cohort of treatment-naïve patients with CHD from Mongolia, a country with a substantial burden of viral hepatitis [13,14].

### Patients and Methods

#### Study population

We identified 51,113 adult individuals (≥18 years) with HBsAg testing at the Liver Center, Ulaanbaatar in Mongolia, between from 1^st^January 2015 - 31^st^ December 2023 **(Figure 1)**. Data were retrieved from an electronic database, comprising medical and investigations records. The Liver Center provides outpatient care for patients with liver diseases, and also screens individuals for viral hepatitis upon self-referral. After exclusions (n=38,056), patients with HBsAg positive were categorized on their virological parameters into two cohorts: 1) HBV/HDV co-infection or CHD, if at least one serum HDV RNA level ≥50 IU/mL (n=3,399), and 2) mono-HBV infection or CHB, with at least one negative anti-HDV and/or HDV RNA test, and no record of anti-HDV or HDV RNA positivity (n=2,556).

**Figure 1:**
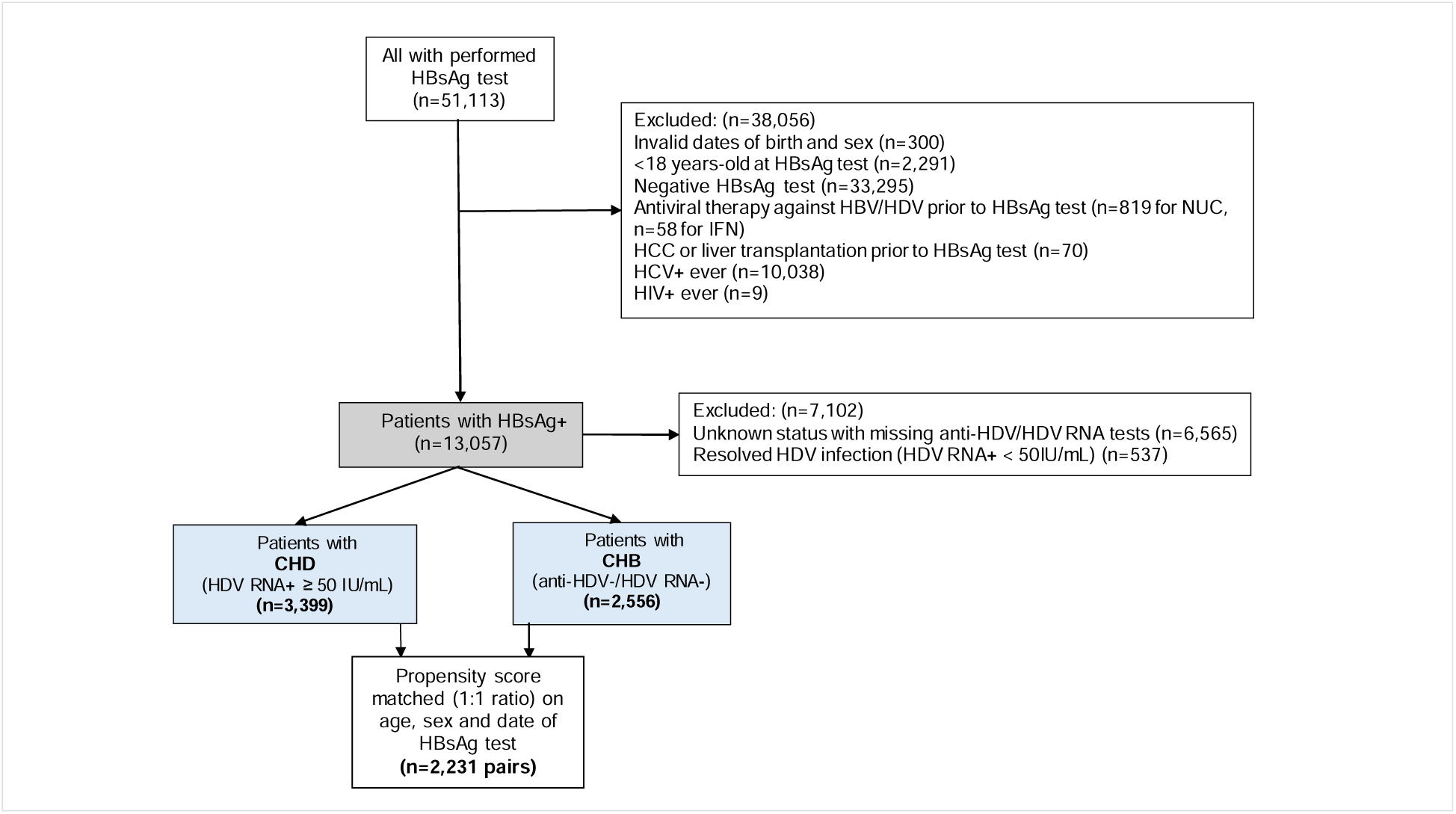
Study flowchart Abbreviations: NUC =nucleo(s)tide analogues; IFN=interferon; HCC= hepatocellular carcinoma; CHD=chronic hepatitis D; CHB=Chronic hepatitis B.

A propensity score matching (PSM) on age, sex, and date of HBsAg test, resulted in 2,231 matched pairs.

All data were anonymized and transferred to Karolinska University Hospital and Karolinska Institutet, Sweden for analysis. Ethical approval was granted by the local ethics review board at the Liver center in Mongolia and by the Ethical Review Authority of Sweden. The study was funded by an ALF grant from Region Stockholm, Sweden.

#### Demographic, biochemical test and fibrosis staging

The baseline date was the first positive HBsAg test date. Demographic data, including date of birth and sex, and medical data such as body mass index (BMI), antiviral treatments and liver-related diagnoses as liver cirrhosis, hepatocellular carcinoma, decompensation events (ascites, variceal bleeding, or encephalopathy) and liver transplantation during the study period of 1^st^January 2015 - 31^st^ December 2023 were retrieved. Biochemical, virological tests and liver stiffness measurements (LSM) were obtained and metabolic risk factors (MRF) were assessed (detailed in the supplements).

#### Statistical analysis

Continuous variables were compared using Student-t or the Mann-Whitney U tests, while categorical variables were compared using the Chi-Square test, or Fisher’s exact test. Due to missing parameters at early visit, a rolling-window of +/-21 days was utilized to capture paired biochemical and virological parameters cross-sectionally within the window period (sTable 1). LSM up to +/-365 days were allowed. The nearest rolling-window to baseline (HBsAg test) with no prior HCC diagnosis, initiation of antiviral treatment, or liver transplantation was considered. Correlations between paired parameters were assessed using Spearman’s correlation coefficient (rho). Coefficient values were interpreted as following: very strong (0.80-1.00), strong (0.60 to <0.80), moderate (0.40 to <0.60), weak (0.20 to <0.40), and very weak (>0.0 to <0.20) [15]. Scatter plots with overlay smooth-lines were used to visualize univariable correlations, representing trends in data. Univariable and multivariable logistic regression analyses were conducted to identify parameters significantly associated with ALT elevation (> upper limit of normal, >ULN), HDV RNA ≥100,000 IU/ml, HBsAg ≥10,000 IU/ml, or presence of LSM ≥15.2 kPa[16]. Results were presented as odds ratio (OR) and 95% confidence intervals (CI). For the multivariable analysis, parameters available within 21 days before and up to 90 days after the baseline date were used. Also, factors associated with CHD were assessed across age groups 18-29, 30-44, 45-59 and ≥60 years of age. Propensity scores matching (PSM) were constructed using logistic regression model with CHD as the outcome, and variables age, sex, and date of HBsAg test as covariates. The model estimated the probability of being CHD per each patient, and the propensity scores were used in 1:1 nearest neighbor matching for age and date of HBsAg and exact matching for sex to balance the groups. After matching, the standardized mean differences (SMD) between CHB and CHD were assessed. All analyses were similarly conducted on the matched pairs.

Heat maps on baseline data of PSM cohorts were used to visualize Spearman’s coefficient of the correlation matrix. Statistical significance was considered when p-values <0.05. Data analyses were executed using R Studio version 3 and IBM SPSS Statistics, version 28.0.1.1.

## Results

### Baseline characteristics of patients with CHD by age groups (*Table 1*, *Figure 2*)

As shown in Table 1, patients aged 18-29 years-old constituted 10.0% of the cohort, men were significantly less prevalent in older age groups compared to women (51.2% in 18-29 years vs 23.5% in ≥60 years, p<0.001). Group of 18-29 years had the highest median ALT 69.3 (44.1-124.0) IU/L, with 15.5% having normal level (p<0.001). To the contrast, GGT level increased with age with elevated levels at 34.6% among 18-29 years reaching 43.7% in older age groups (p<0.001). Young age group also had the highest prevalence of HBeAg+ at 45.0%, higher HBsAg level at 3.9 log_10_ IU/mL and lower median HBV DNA level at 2.1 log_10_ IU/mL compared to older age groups (p<0.001). Of note, HDV RNA level was similar across age groups (p=0.30).

**Figure 2:**
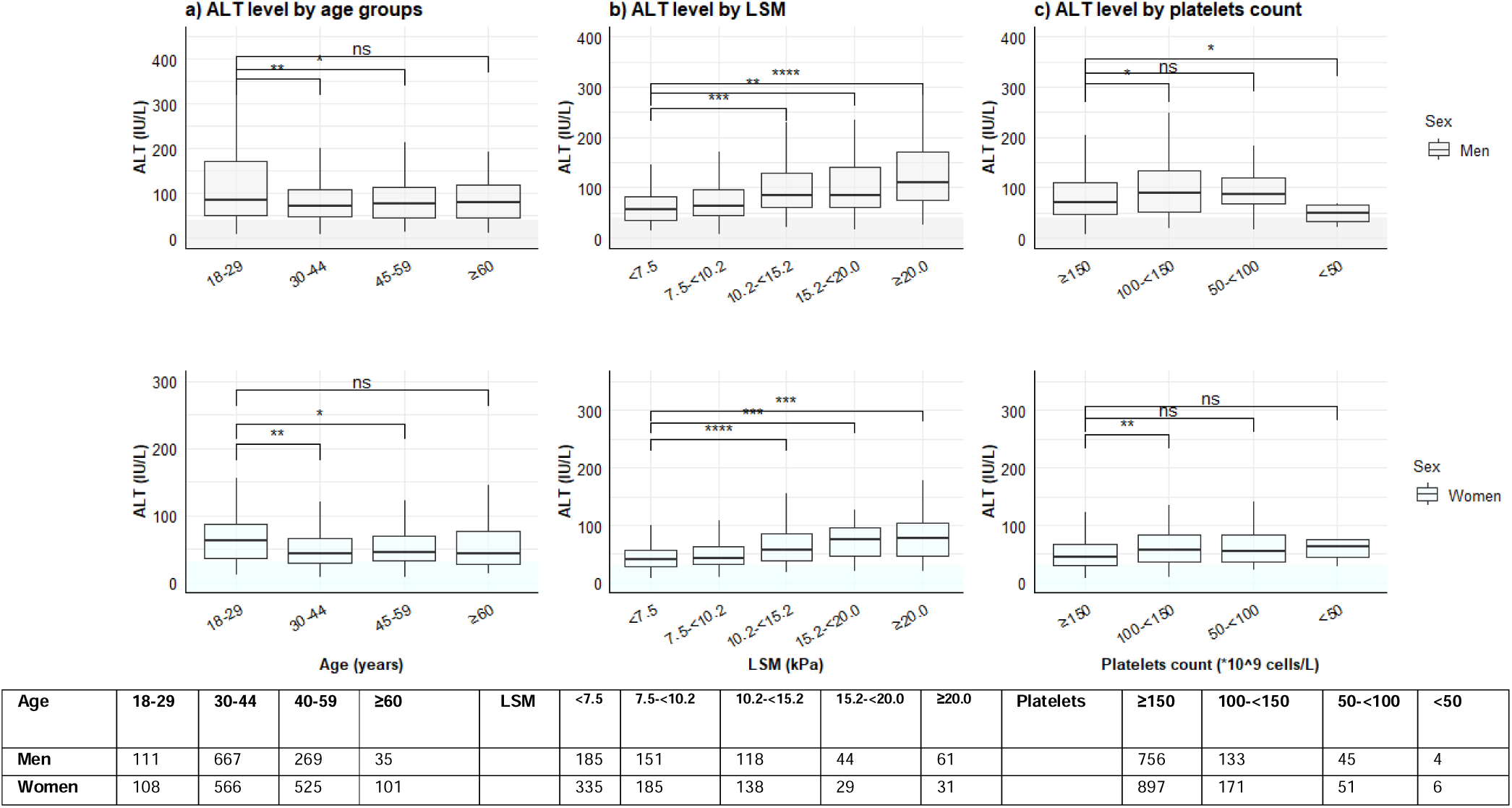
Box plot illustrating ALT levels, subgrouped by a) age, b) liver stiffness measurements and c) platelets counts in men and women with CHD. Abbreviations: ALT=alanine aminotransferase; LSM= liver stiffness measurement. ALT levels on Y-axis, and age categories, liver stiffness and platelets count subgroups are shown on X-axis. Median ALT values are represented by interior bars, with upper and lower borders representing 25^th^ and 75^th^ quartiles, with whiskers of minimum and maximum values. Shaded area mark the upper limit of normal ALT for men (<41 IU/L) and women (<31 IU/L), respectively. Significance was evaluated by T-test. Table shows the number of persons with CHD per category and sex.

**Table 1:**
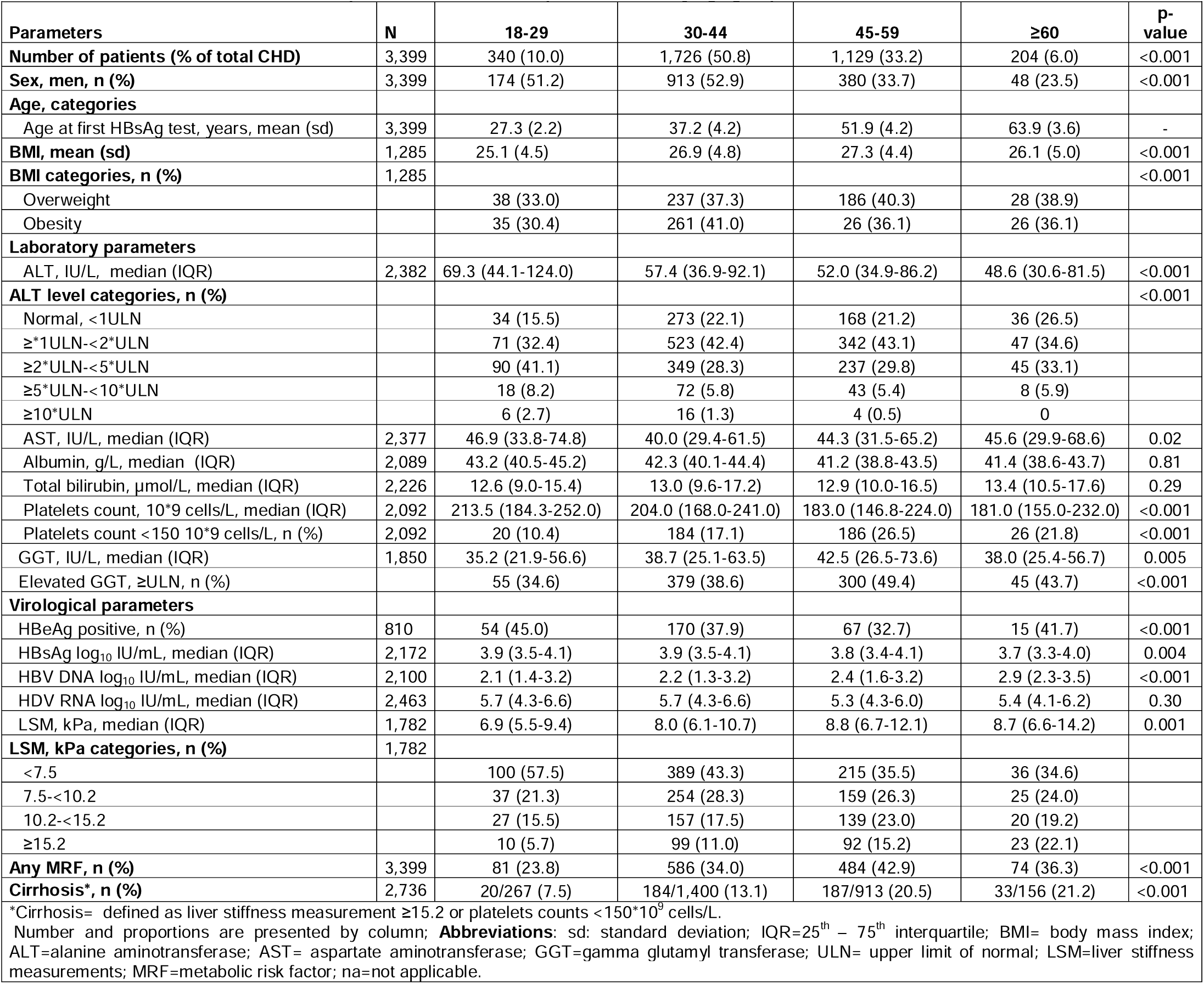
Baseline characteristics of 3,399 patients with chronic hepatitis D (CHD) by age groups.

Figure 2 shows ALT level by age groups in men and women with CHD.

sFigure 1 shows ALT level by age groups, LSM and platelets count in all CHB patients where notably ALT level is similar across age groups.

sTable 2 summarizes baseline characteristics in CHD by sex, and record of cirrhosis.

Briefly, men constituting 44.6% of CHD cohort, were significantly younger and showed higher ALT, AST, GGT, bilirubin levels, LSM as well as more prevalent obesity, MRF and cirrhosis compare to women (also shown in Figure 2). Patients with cirrhosis had higher ALT levels, with only 10.0% having normal ALT compared to 22.9% in those without cirrhosis record (p<0.001).

HDV RNA level ≥100,000 IU/mL was more prevalent patients with cirrhosis (66.1% vs 58.0%, p=0.006), while to the opposite HBsAg ≥10,000 IU/mL was less prevalent in cirrhosis (27.2% vs 35.6%, p=0.005).

### Baseline differences between patients with CHD and CHB (sTable 3)

Compared to CHB, patients with CHD were older (mean age 42.7 vs 36.6 years-old, p<0.001),with less prevalent men (44.6% vs 52.5%, p<0.001).

In CHD, median ALT value was 56.6 (36.5-92.1) IU/L with normal level in 21.5% vs 62.3% in CHB (p<0.001). CHD showed higher AST, GGT, bilirubin, and lower platelets count and LSM (all p<0.05). They showed also higher HBsAg and lower HBV DNA levels, more prevalent MRF (36.0% vs 27.5%) and more cirrhosis record (15.5% vs 3.9%). sTable 4 shows the association of baseline parameters with CHD diagnosis across age groups derived by logistic regression analysis. Age group 18-29 years with CHD demonstrated 8.04-odds of elevated ALT, 10.56-odds of elevated AST, 5.70-odds of low platelets count and 2.89-odds of elevated GGT compared to same age group with CHB. Patients with CHD showed consistently higher odds of HBsAg ≥10,000 IU/mL, more pronounced in older age groups; OR=6.0 in those aged ≥60 years while OR=1.60 in age 18-29 years. Younger group with CHD also showed 3.40-odds of low HBV DNA <2,000 IU/mL, and 4.15-odds of cirrhosis vs peers with CHB.

### Pair-wise correlations of biochemical and virological parameters in CHD and CHB

As shown in (Figure 3), ALT weakly correlated with HDV RNA (rho=0.23, p<0.001) and LSM (rho=0.37, p<0.001), correlated moderately with GGT (rho=0.48, p<0.001), and showed negative weak correlation with platelets count (rho=-0.15, p<0.001). ALT did not correlate with HBV DNA (rho=0.02, p=0.32).

**Figure 3:**
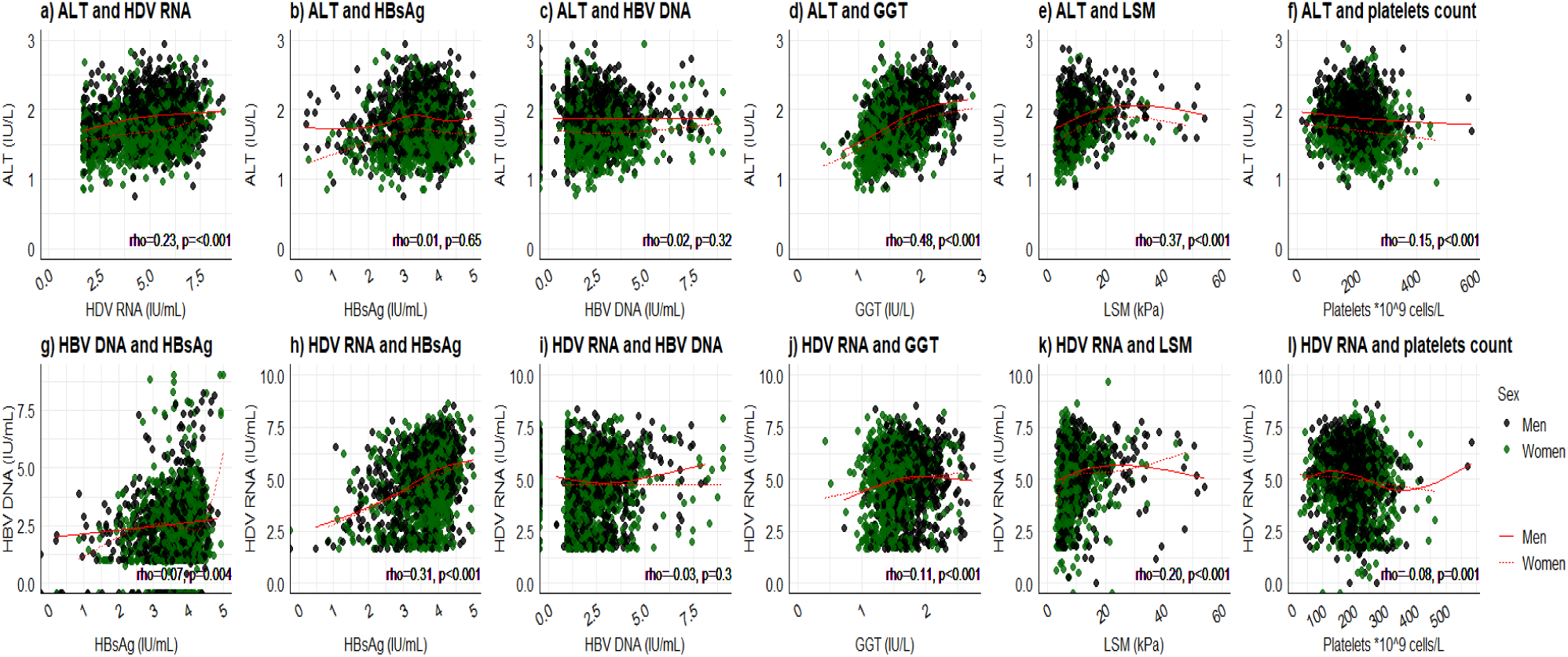
Scatter plots illustrating the association between ALT and HDV RNA levels with other parameters in patients with CHD, subgrouped by sex. A flexible regression line (red) illustrates the direction of the association for each sex. Spearman correlation coefficients (rho) with p-values are shown for the whole group. Abbreviations: ALT=alanine aminotransferase; HDV RNA= hepatitis D virus ribonucleic acid; HBsAg= hepatitis B surface antigen; HBV DNA= hepatitis B virus deoxyribonucleic acid; GGT= gamma glutamyl transferase; LSM=liver stiffness measurement

HDV RNA weakly correlated with HBsAg (rho=0.31, p<0.001), with LSM (rho=0.20, p<0.001), and showed a weak positive correlation with GGT (rho=0.11, p<0.001). No correlation was noted between HDV RNA and HBV DNA levels (rho=0.03, p=0.3). Overall similar correlations were noted in men and women.

HBsAg showed a very weak positive correlation with HBV DNA levels (rho=0.07, p=0.004). HBV DNA and HBsAg levels showed negligible or no correlation with platelet counts and LSM levels.

Pair-wise correlations of parameters in CHD, grouped by cirrhosis are showed in sFigure 2. Overall HDV RNA correlations with ALT, HBsAg, GGT demonstrated similar correlations in non-cirrhosis versus cirrhosis groups.

The correlations in CHB are shown in sFigure 3, where ALT displayed weak correlations with HBsAg (rho=0.16, p<0.001), with HBV DNA (rho=0.19, p<0.001), with platelets (rho=0.18, p<0.001), but stronger correlation with GGT (rho=0.61, p<0.001), and with LSM (rho=0.46, p=0.002). HBV DNA and HBsAg moderately correlated in CHB (rho=0.37, p<0.001).

Pair-wise correlations in CHB, subgrouped by cirrhosis are shown in sFigure 4. Overall stronger correlations between ALT with HBsAg, and GGT in non-cirrhosis versus cirrhosis groups.

**Figure 4:**
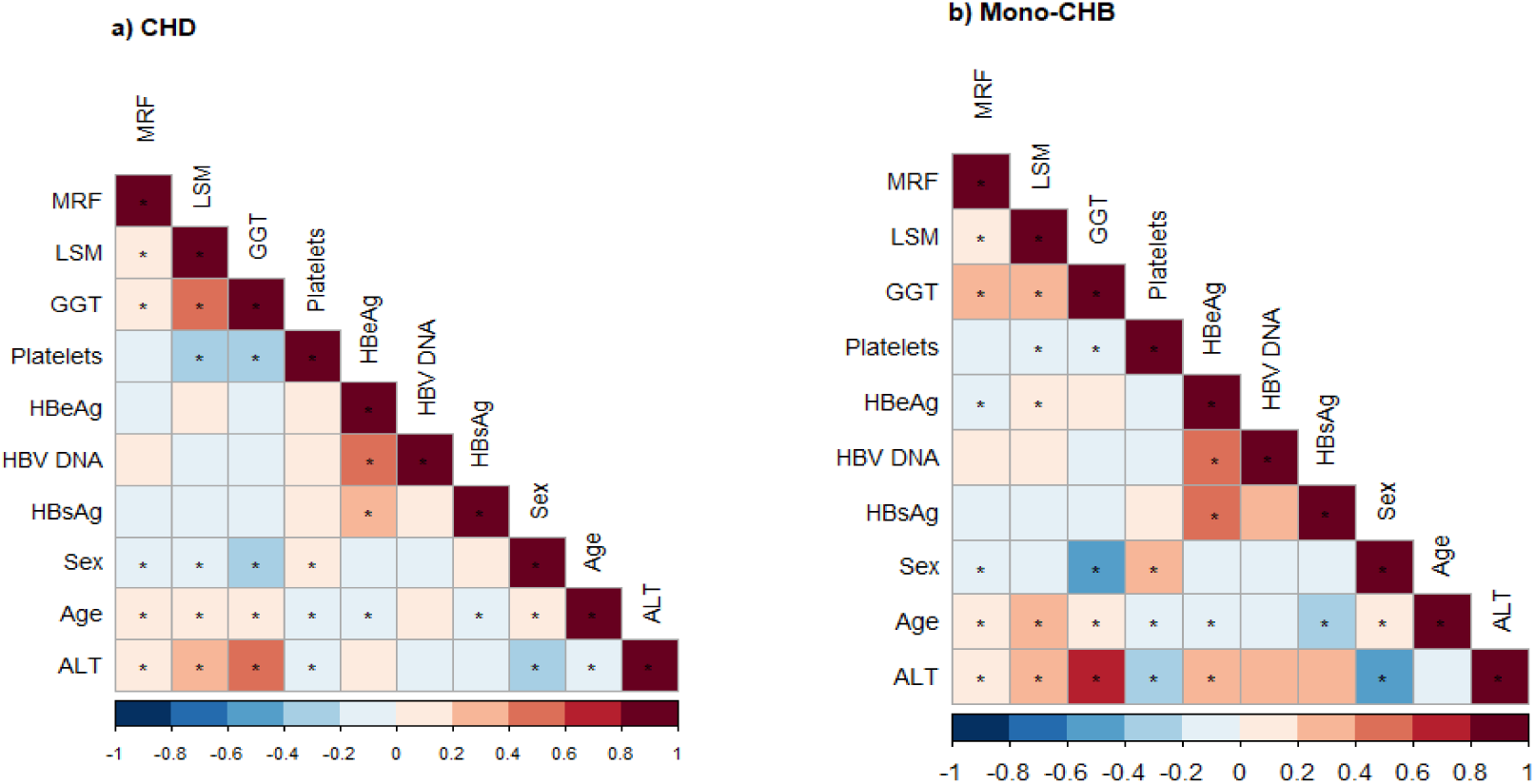
Heatmaps showing the pairwise correlations in 2,231 propensity score-matched pairs of patients with chronic hepatitis D (CHD) and chronic hepatitis B (CHB). The correlation coefficient values are represented by a color gradient, with significant correlations marked by an asterisk (*). Abbreviations: MRF=metabolic risk factor; ALT=alanine aminotransferase; HBsAg= hepatitis B surface antigen; HBV DNA= hepatitis B virus deoxyribonucleic acid; GGT= gamma glutamyl transferase; LSM=liver stiffness measurement. Sex, blue=association with men. Parameters on continuous scale, except HBeAg, sex, and MRF.

### Factors associated with elevated ALT, high HDV RNA-, HBsAg levels or LSM ≥15.2 kPa in CHD

Factors associated with elevated ALT were shown in Table 2; where age <30 years (aOR=2.86), elevated GGT level (aOR=3.96), and HDV RNA ≥100,000 IU/mL (aOR=2.48) remained significant in multivariable analysis.

**Table 2:**
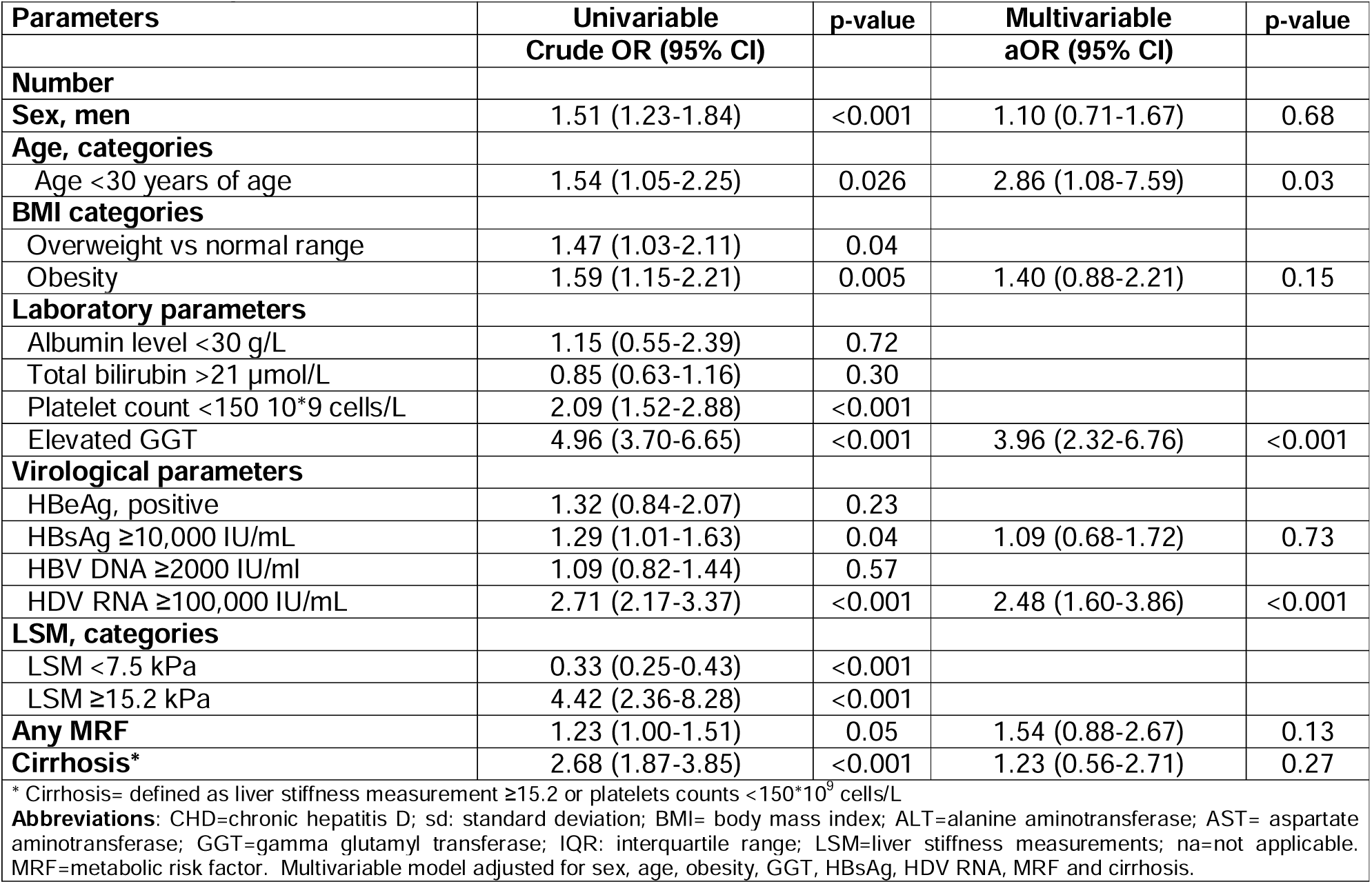
Parameters associated with elevated ALT level in patients with CHD (n=3,399) in univariable and multivariable logistic regression models. Crude Odds ratio (OR) and adjusted OR (95% confidence intervals, CI) are presented.

sTable 5 show that for LSM ≥15.2 kPa, the following factors remained significant: male sex (aOR=1.97), elevated bilirubin levels (aOR=2.94), elevated GGT levels (aOR=5.29), platelet counts <150.0 *10^9^ cells/L (aOR=5.73), HDV RNA ≥100,000 IU/mL (aOR=2.38) and the presence of MRF (aOR=2.00).

In sTable 6 multivariable analyses showed that only HBsAg ≥10,000 IU/mL remained a significant factor with HDV RNA level ≥100,000 IU/mL. sTable 7 shows that elevated GGT (aOR=0.48), HBeAg+ (aOR=1.98) and HDV RNA level ≥100,000 IU/mL (aOR=2.79) remained significant factors associated with HBsAg level ≥10,000 IU/mL, in multivariable analyses. Of note, women showed a positive trend with (aOR=1.64) but not reaching statistical significance (p=0.06)

### Comparison between propensity-score matched CHD and CHB (sTable 8)

*As previously shown due to the significant different age and sex distribution between CHD and CHB cohorts, we performed a* propensity-score matching (on age, sex and date for HBsAg test) which yielded 2,231 pairs of CHD and CHB with similar age and sex distribution shown in sTable 8. The SMD for propensity score (distance) was 0.06, for sex was 0.0001, for age 0.06 and for date of HBsAg was 0.002. All SMDs were <0.1, indicating excellent balance across the matched covariates (results not tabulated).

In sTable 9, factors remaining associated with CHD in PSM cohorts were elevated ALT (aOR=2.58), elevated AST (aOR=5.49), low platelets count (aOR=4.27), HBeAg+ (aOR=2.46), and low HBV DNA replication (aOR=6.18).

ALT elevation was especially higher in individuals 18-29 years of age, where they had 4.80-odds of elevated ALT, 2.76-odds of elevated GGT, 12.3-odds of elevated AST and 5.1-odds of cirrhosis compared to PSM peers with CHB of the same age group (results not shown in Table).

Heatmaps show the summary of univariable correlations in PSM cohorts of CHD and CHB separately (Figure 4).

## Discussion

In this cohort study of 3,399 treatment-naïve Asian patients with CHD, compared to CHB, we identified following key findings: 1. Younger CHD patients, aged 18-<30 years, exhibited significantly higher levels of ALT, AST and GGT and more frequent advanced fibrosis than age-matched CHB patients; 2. Age <30 years and ALT elevation was associated with, elevated GGT and HDV RNA ≥100,000 IU/mL to the contrast of CHB. To our knowledge, this is the largest study to date, examining the prevalence and patterns of ALT elevation in CHD, compared to a matched CHB cohort, offering new insights into the distinct features of HDV infection, with potential implications for timing and strategies of HDV treatment.

Consistent with prior studies, patients with CHD showed higher transaminases and advanced fibrosis compared to CHB, alongside a higher prevalence of HBeAg positivity [6,17,18]. Our novel finding of a more frequent and pronounced ALT elevation in young adults with CHD suggests a more active or prolonged phase of immune response targeting infected hepatocytes than previously known compared to HBV-monoinfection [19]. Earlier studies lacked sufficient young adults data and were not powered for age-specific analyses [6,17,18]. This severe hepatic inflammation in young adults with CHD, may reflect an elicited immune response to HBV/HDV co-infection when entering early adult life, or that HDV superinfection occurred in adolescence or early adulthood when becoming sexually active in a high-endemic region. The latter hypothesis is consistent with Mongol data estimating HDV prevalence among school children with HBsAg+ at 13.6%, while reaching 60-80% in adults with HBsAg+ implying that later HDV superinfection is more predominant [20,21].

Unlike CHD with poorly characterized phases in its natural course, the phases of CHB have been defined as following: HBeAg-positive chronic HBV infection (immune tolerant phase), HBeAg-positive chronic hepatitis (immune active phase), HBeAg-negative chronic HBV infection (inactive carrier phase), HBeAg-negative chronic hepatitis (immune escape phase) and HBsAg-negative phase (resolved HBV infection phase) [22]. Individuals acquiring mono-HBV in adolescence or adulthood often bypass or shorten “HBeAg+ chronic infection” phase, which typically extends for decades in perinatal or early childhood infected patients [23]. This transition into chronic hepatitis, with a shortened/or without an immune-tolerant phase, suggests a more accelerated disease course, particularly among older-age HDV infections [24]. Delayed HBeAg seroconversion, typically occurring at age 30-35 years in HBV-monoinfection is associated with increased risk of cirrhosis [25]. In the present analysis, CHD patients aged 30-44, and 40-55 years had 1.6-respectively 2.1-fold higher odds of HBeAg positivity compared to CHB peers, indicating delayed seroconversion [26]. Nevertheless, this prevalence of HBeAg+ in middle and older age individuals with CHD in our analysis is higher compared to western cohorts with ∼15% HBeAg+ at diagnosis [6,11]. However, extrapolating HBV phases in CHD remains challenging, with most patients fitting into *HBeAg negative chronic hepatitis* characterized by HBeAg-, elevated ALT but suppressed HBV DNA levels.

Persistent HDV RNA replication and elevated ALT levels are associated with accelerated risk of cirrhosis and promote carcinogenesis, by processes of necroinflammation, fibrosis accumulating oxidative stress, and DNA damage [27]. ALT levels declined in the late stages of advanced fibrosis and cirrhosis, consistent with previous studies, with a negative correlation between ALT and platelets count [27,28]. However, normal ALT does not exclude ongoing low-grade hepatic inflammation, also seen in CHB, warranting cautious interpretation of normal ALT levels in CHD [29,30]. Our findings support considering treating patients with detectable HDV RNA regardless of age and disease phase, considering the severe necro-inflammation and fibrosis early in life; to prevent later severe liver-complications. This align with recent EASL and WHO guidelines in which all patients should be considered for HDV therapy [7,31]. Similarly, a recent simulated cost-effectiveness analysis concluded that early treatment (F0 fibrosis of CHD) significantly reduced liver-related complications, including HCC and was cost-effective [32]. Currently the access of the recently approved HDV drug of bulevirtide is restricted to advanced fibrosis/cirrhosis in some countries with available therapy [33]. EASL guidelines recommends though that all patients with CHD should be considered for antiviral therapy [7].

In agreement with prior studies *(mostly in individuals under therapy)*, HDV RNA and HBsAg levels correlated positively, albeit weak (rho=0.33, p<0.001). HBsAg levels did not correlate with LSM or platelets count, questioning its reliability as a potential marker of CHD progression. This contrasts to other published studies with limited sample sizes, in which HBsAg level was associated with histological activity, particularly in non-cirrhotic cases [34]. Also, HBV DNA replication was associated with worsened outcome of CHD, regardless of HDV RNA level [35]. This highlights the complex interplay between HDV, HBV and host immunity [36].

In a Spanish study, 25% of CHD patients achieved a spontaneous ≥2log HDV RNA decline over 5 years, associated with corresponding declines in ALT, HBV DNA and HBsAg levels [37]. Our cross-sectional data showed that HDV RNA levels did not vary across age groups, suggesting no natural decline, although a longitudinal analysis would better answer this question.

ALT normalization, a surrogate for improved clinical outcomes in CHD under treatment, occurs in 30-40% with peg-IFN monotherapy [38], and in 50-60% with recently introduced bulevirtide monotherapy [39]. In phase 3 clinical trial of bulevirtide, a discrepancy of biochemical response of ALT (normalization of ALT from elevated level at baseline) and virological response of HDV RNA (≥ 2 log decrease from baseline) was observed during treatment in some patients, with the underlying reason remaining unknown [39]. Patients with HDV RNA decline but failure to reach ALT normalization are deemed partial responders. Other confounding factors such as comorbidities of MRF (36% in the current cohort) should be considered when assessing ALT responses [7]. In that context, GGT might be a useful marker of liver disease as it correlated with ALT (rho=0.49, p<0.001), and with cirrhosis agreeing with prior studies, and showed the strongest correlation with HDV RNA levels in the present analysis [40].

We acknowledge several limitations in the present study. The cross-section design with significant correlations cannot establish causal relationship. However, analysis in a large treatment-naïve cohort may help to gain deeper understanding of CHD, define clinical phenotypes, and generate hypothesis for experimental and longitudinal studies. The mode and age at infection were not directly ascertainable; however, HBV in endemic regions is typically acquired perinatally or during early childhood, while HDV superinfection likely occurs later.

Other granular data on HDV or HBV genotype, socioeconomic, alcohol consumption, health behaviors, therapies except for anti-HBV/HDV therapies were not assessed. Previous studies have though indicated that HDV genotype 1 and HBV genotype D predominate in Mongolia [13]. Hence, our data may not be generalizable to other genotypes. The use of liver stiffness and platelets might misclassify cirrhosis, however we used recently biopsy-proven proposed cut-offs to classify the fibrosis stages [16]. The study setting, reflective of real-world practice in resource-limited regions, caused missing data and introduced potential selection bias towards patients with more symptomatic disease, and those who can afford tests. For this we conducted a complete case analysis, and rolling-window to maximize the usage of available data. Our data include a population of only Asian ethnicity from one country, but a large and rather homogenous cohort in our study could serve as a model to understand ALT elevation and associations with other factors in CHD, with statistical power and possibility to adjust for confounders by this approach.

In conclusion, significant liver inflammation, as indicated by elevated ALT levels, distinguishes CHD from CHB in young adults. Our findings support early HDV treatment in patients with detectable HDV RNA to halt CHD progression and reduce HCC risk. Further studies are needed to clarify the role of ALT, the virological and immunological interactions driving CHD pathogenesis.

## Data Availability

All data produced in the present work are contained in the manuscript

## Abbreviations

CHD: chronic hepatitis D
ALT: alanine aminotransferase
MRF: metabolic risk factor

## Authors contributions

All authors contributed to the study conception and design. Material preparation, data collection [Ganbolor Jargalsaikhan], [Sanjaasuren Enkhtaivan], [Habiba Kamal] and analysis were performed by [Habiba Kamal] and [Daniel Bruce]. The first draft of the manuscript was written by [Habiba Kamal] and [Soo Aleman] and all authors commented on previous versions of the manuscript. All authors read and approved the final manuscript.

## Acknowledgements

Staff involved in the study, and people living with chronic hepatitis B and D and their care givers.

