## Supplementary material for "Prevalence and risk factors of elevated ALT in 3,399 treatment-naïve HBV/HDV co-infected patients: A comparison with propensity score-matched HBV mono-infected patients": Supplemetal tables and figures

**Supplementary document:**

**Methods:**

*Demographic, biochemical test and fibrosis staging*

Body mass index (BMI) was calculated as weight in kilogram divided by height in meters squared (kg/m^2^). The following cut-offs were used for BMI categories in accordance with Asian population standards: underweight ( <18.5 kg/m^2^), normal weight (18.5 -22.9 kg/m^2^), overweight (23.0- 27.4 kg/m^2^) and obesity (≥ 27.5 kg/m^2^) [1].

The upper limit of normal (ULN) for biochemical tests, as per local laboratory routines, were defined as following: ALT of 41 IU/L for men and 31 IU/L for women, AST of 35 IU/L for men and 31 IU/L for women, GGT of 55 IU/L for men and 38 IU/L for women, serum triglyceride of 1.7 mmol/L, and serum low-density lipoproteins of 2.6 mmol/L. The lower limit of normal (LLN) for serum high density lipoproteins was 1.0 mmol/L for men and 1.3 mmol/L for women.

Metabolic risk factor were defined, as presence of at least one of following conditions: overweight or obesity, diabetes mellitus (either a documented diagnosis or fasting serum glucose ≥7.0 mmol/L on two occasions), dyslipidemia, hypertension and/or diagnosis of fatty liver disease [2]. Events of HCC, decompensation events (defined as ascites, variceal bleeding, or encephalopathy), or liver transplantation were retrieved from medical records, as well as from ultrasound or gastroscopy investigation records where appropriate.

For CHD, the LSM cut-offs used to define advanced/F3 fibrosis was ≥10.2−<15.2 kPa, while ≥15.2 kPa for F4 fibrosis [3]. The corresponding LSM cut-off levels for CHB were ≥9.0-<12.5 kPa for F3 and ≥12.5 kPa for F4 fibrosis [4]. LSM values with a success rate of ≥90% and the interquartile range (IQR) of ≤30% were considered reliable and used [4]. Since LSM levels may be affected by elevated transaminases due to liver necroinflammation [5], sensitivity analyses using platelets count as an alternative marker for advanced fibrosis/cirrhosis were performed. For cirrhosis diagnosis, criteria of either a LSM value of ≥15.2 for CHD or ≥12.5 kPa for CHB, or platelet counts of <150*10^9^ cells/L was utilized [6].

*Serological and virological tests*

HBsAg was tested with either rapid diagnostic test (CTK Co.LtD, USA), or quantitative HBsAg test (Sysmex Co.Ltd, Japan) at baseline. Anti-HCV and anti-HIV were qualitatively tested by rapid diagnostic test (CTK Co.LtD, USA). Quantitative HBsAg was estimated by HISCL-5000 fully automated chemiluminescence analyzer (Sysmex Co.Ltd, Japan) and for HBeAg status (CLIA, Sysmex, Japan, lower level of detection (LOD) of 1 COI). HCV RNA was quantified using real time reverse transcriptase polymerase chain reaction with LOD of 10 IU/mL (GeneXpert, Cepheid, USA). Anti-HDV was analyzed using the Wantai HDV-IgG ELISA (Wantai Biopharmaceutical Co. Ltd, China), with a lower limit of detection (LoD) of <1 COI. HDV RNA levels were quantified, using a Bioactiva Diagnostica extraction kit (Bioactiva Diagnostica, Germany) and a Bio-Rad amplification kit (Bio-Rad Laboratories, USA) with a lower limit of quantification (LoQ) of <50 IU/mL. HBV DNA levels were analyzed using the GeneXpert system (Cepheid, USA), with an LoQ of <10 IU/mL.

**Supplementary figures:**

**sFigure 1**: Box plot illustrating ALT levels subgrouped by age, liver stiffness measurements (LSM), and platelets count CHB.

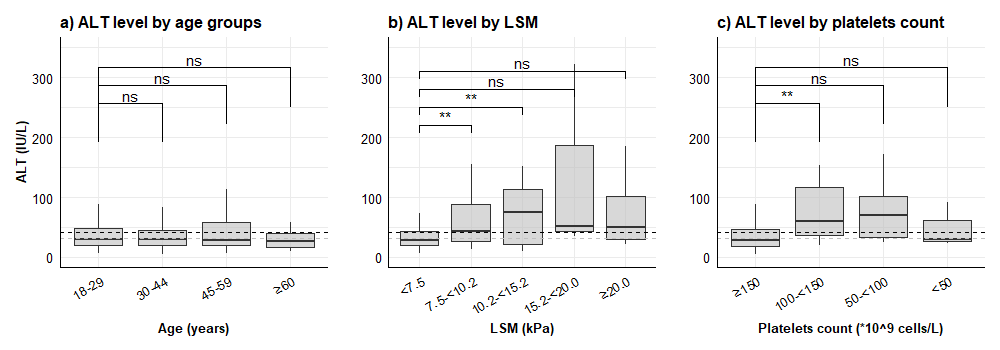

| **Age** | **18-29** | **30-44** | **40-59** | **≥60** | **LSM** | **<7.5** | **7.5-<10.2** | **10.2-<15.2** | **15.2-<20.0** | **≥20.0** | **Platelets** | **≥150** | **100-<150** | **50-<100** | **<50** |
| --- | --- | --- | --- | --- | --- | --- | --- | --- | --- | --- | --- | --- | --- | --- | --- |
| **Total** | 384 | 890 | 233 | 46 |  | 367 | 47 | 17 | 4 | 10 |  | 1191 | 27 | 10 | 3 |

ALT level on Y-axis, age categories, liver stiffness and platelets count subgroups are shown on X-axis. Median ALT values are represented by interior bars, upper and lower borders represent 25^th^ and 75^th^ quartiles and whiskers represent minimum and maximum values. Black and grey dashed lines mark the upper limit of normal for men (<41 IU/L) and women (<31 IU/L), respectively. ALT elevation was similar across age groups in CHB. Patients with LSM <7.5 kPa had significantly lower ALT than higher LSM groups until <15.0 kPa, then similar ALT level could be seen thereafter. Only platelets count 100-150.0 cells*10^9^/L had significantly higher ALT level than those with ≥150.0 (p<0.001). Significance was evaluated by T-test. Table shows the number of persons with CHB per category and sex.

**sFigure 2:** Scatter dot plots illustrating the association of ALT and HDV RNA levels with other parameters in CHD, subgrouped by cirrhosis*

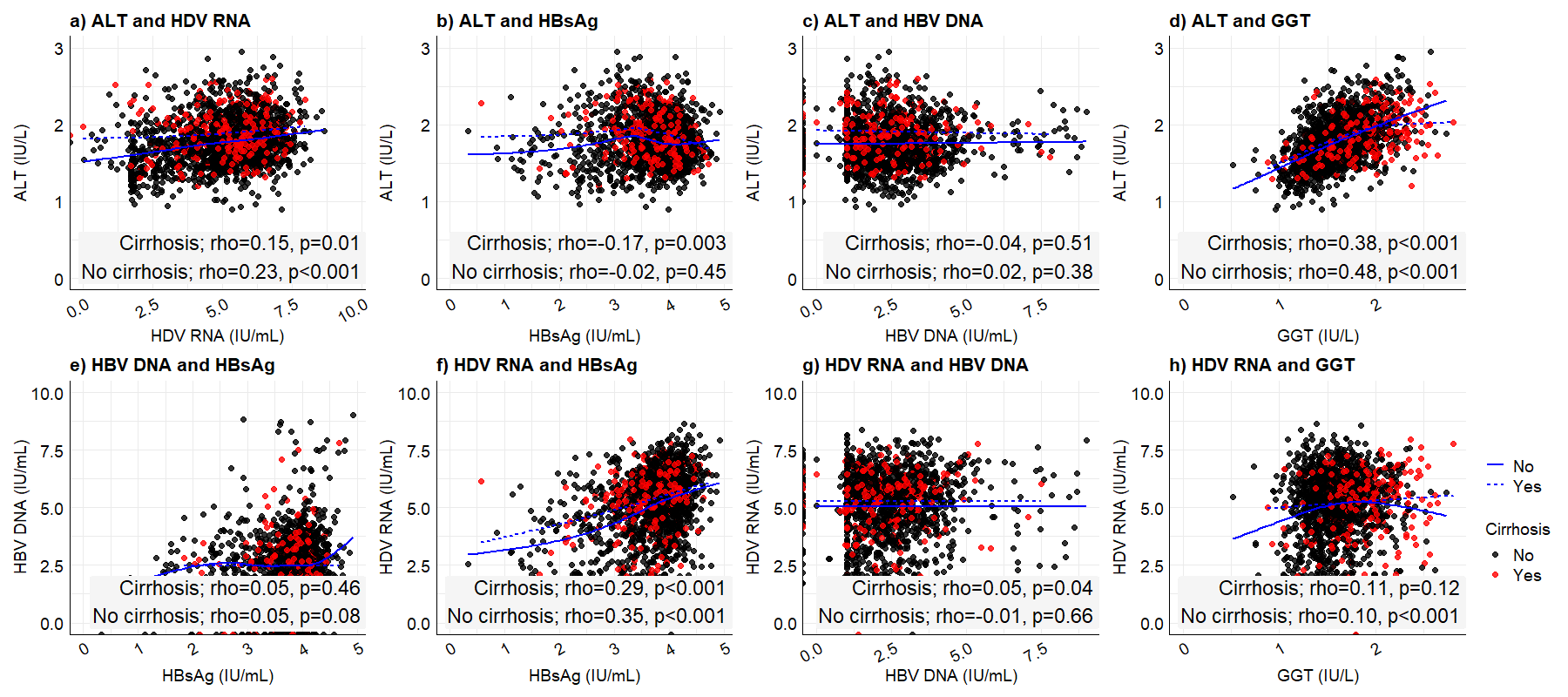

*Cirrhosis is defined as LSM ≥15.2 or platelets count < 150.0*10^9^ cells/L. The pairwise correlation of virological and laboratory parameters at baseline was evaluated using the Spearman correlation coefficient (rho, p-value) and flexible regression line (blue) to illustrate the direction of the association. Abbreviations: ALT=alanine aminotransferase; HDV RNA= hepatitis D virus ribonucleic acid; HBsAg= hepatitis B surface antigen; HBV DNA= hepatitis B virus deoxyribonucleic acid; GGT= gamma glutamyl transferase

**sFigure 3**: Scatter dot plots illustrating the association between ALT and HBV DNA levels with other parameters in CHB subgrouped by sex.

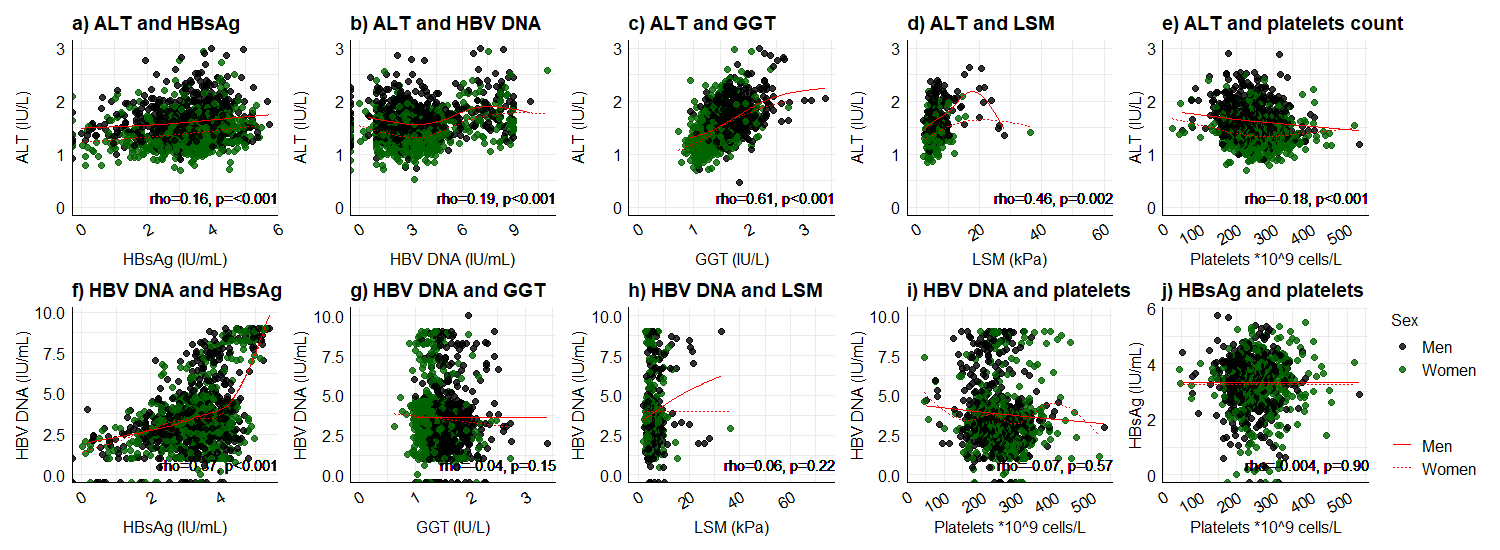

The pairwise correlation of virological and laboratory parameters within 21 days was evaluated using the Spearman correlation coefficient (rho, p-value) and the direction of the association is illustrated as a red line. ALT positively correlated HBsAg, HBV DNA, GGT, LSM and negatively correlated with platelets count (all weak correlations). HBV DNA positively correlated with HBsAg (moderate correlation). GGT, LSM and platelets counts did not correlate with HBV DNA levels, nor with HBsAg levels. ALT=alanine aminotransferase; HDV RNA= hepatitis D virus ribonucleic acid; HBsAg= hepatitis B surface antigen; HBV DNA= hepatitis B virus deoxyribonucleic acid; GGT= gamma glutamyl transferase; LSM=liver stiffness measurement

**sFigure 4**: Scatter plot illustrating the association between ALT and HBV DNA levels with other parameters in CHB subgrouped by cirrhosis*

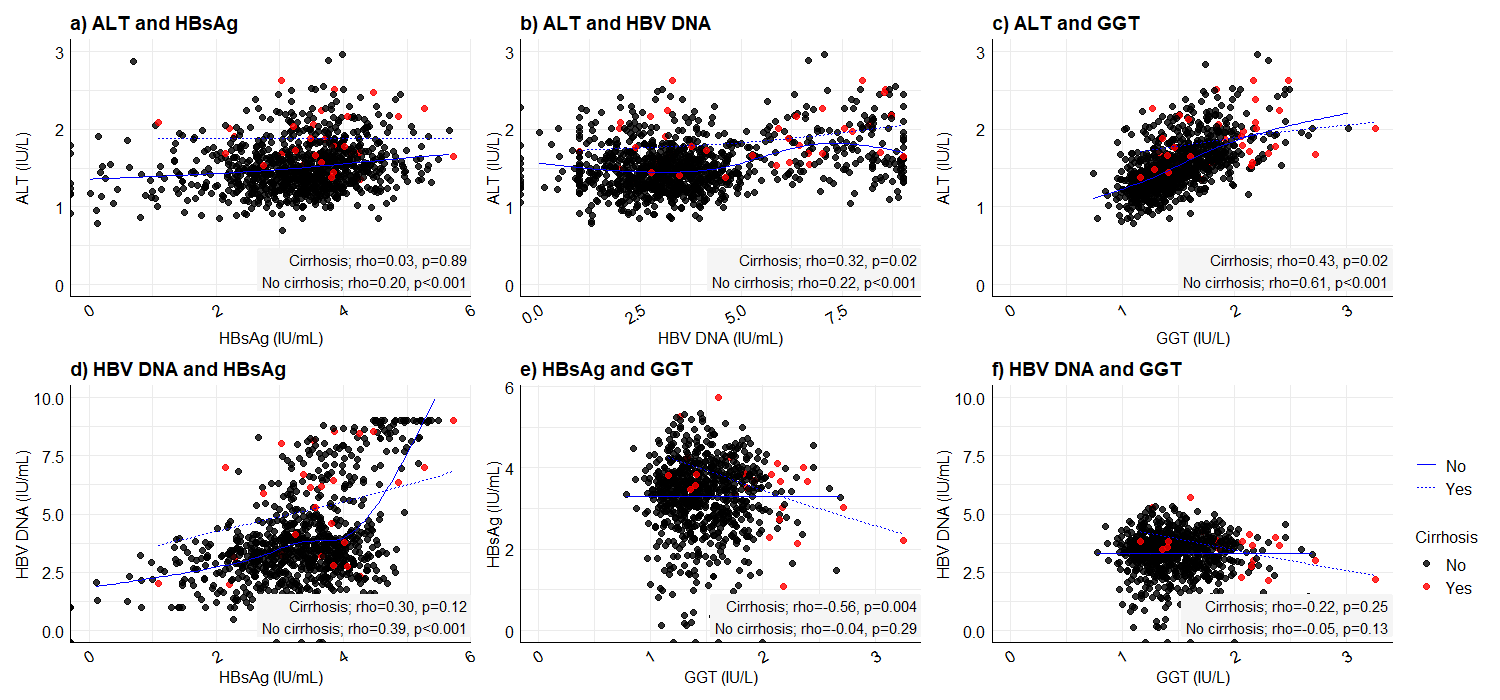

Overall, in no cirrhosis stronger correlations could be noted between ALT and HBsAg, HBV DNA, and between HBV DNA with HBsAg (moderate correlation) and HBsAg and GGT (negative moderate correlation). No correlation between HBV DNA with GGT regardless of cirrhosis. *Defined as LSM≥12.5 or platelets count <150.0 *10^9^cells/L. ALT=alanine aminotransferase; HDV RNA= hepatitis D virus ribonucleic acid; HBsAg= hepatitis B surface antigen; HBV DNA= hepatitis B virus deoxyribonucleic acid; GGT= gamma glutamyl transferase

| sTable 1: Number of unique observations used in correlation analyses | | | | | | | |
| --- | --- | --- | --- | --- | --- | --- | --- |
| Rolling-window variables | ALT | GGT | HDV RNA | HBsAg | HBV DNA | Platelets | LSM |
| ALT | 3,358 | 2,509 | 2,094 | 2,578 | 1,891 | 2,063 | 1,277 |
| GGT | 2,509 | 3,281 | 1,581 | 1,606 | 1,418 | 1,626 | 1,024 |
| HDV RNA | 2,094 | 1,581 | 3,399 | 1,703 | 1,269 | 1,802 | 1,211 |
| HBsAg | 2,578 | 1,606 | 1,703 | 3,358 | 1,397 | 1,759 | 1,137 |
| HBV DNA | 1,891 | 1,418 | 1,269 | 1,397 | 2,326 | 1,557 | 1,121 |
| Platelets | 2,063 | 1,626 | 1,802 | 1,759 | 1,557 | 2,092 | 1,138 |
| LSM | 1,277 | 1,024 | 1,211 | 1,137 | 1,121 | 1,138 | 1,782 |
| Abbreviations: ALT= alanine aminotransferase; GGT=gamma glutamyl transferase; HDV= hepatitis D virus; HBsAg=hepatitis B surface antigen; LSM=liver stiffness measurement. | | | | | | | |

| **sTable 2: Baseline characteristics of patients with chronic hepatitis D (CHD), subgrouped by sex and by cirrhosis** | | | | | | | |
| --- | --- | --- | --- | --- | --- | --- | --- |
|  | **All** | **Women** | **Men** | ***p-***  ***value*** | **No cirrhosis** | **Cirrhosis*** | ***p-***  ***value*** |
| **Parameters** | 3,399 (57.1) | 1,884 (55.4) | 1,515 (44.6) | *<0.001* | 2,312 (84.5) | 424 (15.5) | *<0.001* |
| **Sex, men** | 1,515 (44.6) | na | 1,515 (100) |  | 987 (42.7) | 227 (53.5) | *<0.001* |
| Age at first HBsAg test, years, mean (sd) | 42.7 (10.5) | 44.6 (11.0) | 40.3 (9.4) | *<0.001* | 42.1 (10.4) | 45.6 (10.0) | *<0.001* |
| BMI, mean (sd) | 26.8 (4.7) | 26.4 (4.8) | 27.3 (4.6) | *0.001* | 26.5 (4.5) | 28.9 (5.1) | *<0.001* |
| **BMI categories,** n (%) | *1,285* | *705* | *580* |  | *977* | *171* |  |
| Overweight | 489 (38.1) | 273 (38.7) | 216 (37.2) |  | 375 (38.4) | 55 (32.2) |  |
| Obesity | 528 (41.1) | 264 (37.4) | 264 (45.5) |  | 375 (38.4) | 100 (58.5) |  |
| **Laboratory parameters** |  |  |  |  |  |  |  |
| ALT, IU/L, median (IQR) | 56.6 (36.5-92.1) | 45.8 (31.2-71.9) | 73.0 (47.7-118.1) | *<0.001* | 53.6 (35.0-87.0) | 78.3 (48.5-115.8) | *<0.001* |
| **ALT level categories,** n (%) | *2,382* | *1,300* | *1,082* |  | *1,851* | *361* |  |
| Normal ALT level, <1ULN | 511 (21.5) | 319 (24.5) | 192 (17.7) | *<0.001* | 424 (22.9) | 36 (10.0) |  |
| ≥*1ULN-<2*ULN | 983 (41.3) | 552 (42.5) | 431 (39.8) | *<0.001* | 786 (42.5) | 132 (36.6) |  |
| ≥2*ULN-<5*ULN | 721 (30.3) | 360 (27.7) | 361 (33.4) | *<0.001* | 526 (28.4) | 154 (42.7) |  |
| ≥5*ULN-<10*ULN | 141 (5.9) | 55 (4.2) | 86 (7.9) | *<0.001* | 91 (4.9) | 39 (10.8) |  |
| ≥10*ULN | 26 (1.1) | 14 (1.1) | 12 (1.1) | *0.2* | 24 (1.3) | 0 |  |
| AST, IU/L, median (IQR) | 42.1 (30.4-63.9) | 39.9 (29.6-57.6) | 45.9 (31.8-72.8) | *<0.001* | 39.8 (29.7-58.4) | 63.0 (43.5-94.5) | *<0.001* |
| Albumin, g/L, median (IQR) | 42.0 (39.6-44.1) | 41.4 (39.1-43.4) | 42.8 (40.3-45.0) | *0.37* | 41.9 (40.0-44.1) | 39.4 (36.3-42.2) | *0.62* |
| Total bilirubin, µmol/L, median (IQR) | 12.9 (9.6-16.8) | 12.0 (8.8-15.5) | 14.1 (10.7-18.3) | *<0.001* | 12.5 (9.3-16.0) | 16.0 (11.3-20.9) | *<0.001* |
| Platelets count, 10*9 cells/L, median (IQR) | 198.7 (159.0-236.0) | 199.8 (159.0-240.0) | 198.0 (159.3-231.6) | *0.2* | 207.5 (177.0-244.0) | 125.0 (97.4-147.0) | *<0.001* |
| Platelets count <150 10*9 cells/L | 416/2,092 (19.9) | 232/1,138 (20.4) | 184/954 (19.3) | 0.53 | 133/1,738 (7.7) | 283/354 (79.9) | *<0.001* |
| GGT, IU/L | 39.6 (25.4-65.0) | 31.8 (21.3-49.9) | 51.7 (34.1-86.0) | *<0.001* | 36.3 (24.0-56.2) | 75.3 (42.5-122.8) | *<0.001* |
| Elevated GGT,≥*1ULN, n (%) | 779/1,850 (42.1) | 382/984 (38.8) | 397/866 (45.8) | 0.002 | 516/1,447 (35.7) | 203/280 (72.5) | <0.001 |
| **Virological parameters** |  |  |  |  |  |  |  |
| HbeAg positive, n (%) | 306/810 (37.8) | 160/463 (34.6) | 146/347 (42.1) | *0.029* | 230/587 (39.2) | 32/84 (38.1) | *0.85* |
| HBsAg log_10_ IU/mL | 3.8 (3.4-4.1) | 3.9 (3.5-4.1) | 3.8 (3.4-4.1) | *0.007* | 3.8 (3.4-4.1) | 3.8 (3.5-4.0) | *0.88* |
| HBsAg ≥10,000 IU/mL, n (%) | 722/2,172 (33.2) | 424/1,188 (35.7) | 298/984 (30.3) | *0.008* | 578/1,624 (35.6) | 82/302 (27.2) | *0.005* |
| HBV DNA log_10_ IU/mL | 2.2 (1.3-3.2) | 2.2 (1.3-3.1) | 2.4 (1.4-3.3) | *0.003* | 2.3 (1.4-3.1) | 2.2 (1.3-3.1) | *0.104* |
| HDV RNA log_10_ IU/mL | 5.4 (4.3-6.2) | 5.3 (4.1-6.1) | 5.5 (4.4-6.2) | *0.01* | 5.3 (4.1-6.1) | 5.4 (4.8-6.2) | *0.014* |
| HDV RNA ≥100,000 IU/mL, n (%) | 1471/2,463 (59.7) | 775/1,346 (57.6) | 696/1,117 (62.3) | *0.02* | 1,000/1,725 (58.0) | 220/333 (66.1) | *0.006* |
| LSM | 8.2 (6.1-11.4) | 7.7 (5.9-10.4) | 8.8 (6.6-12.5) | *<0.001* | 7.7 (6.0-9.9) | 20.5 (17.1-26.8) | *<0.001* |
| **LSM, categories,** n (%) | *1,782* | *1,026* | *756* |  | *1,558* | *224* |  |
| <7.5 kPa | 740 (41.5) | 482 (47.0) | 258 (34.1) |  | 740 (47.5) | 0 |  |
| 7.5-<10.2 kPa | 475 (26.7) | 271 (26.4) | 204 (27.0) |  | 475 (30.5) | 0 |  |
| 10.2-<15.2 kPa | 343 (19.2) | 182 (17.7) | 161 (21.3) |  | 343 (22.0) | 0 |  |
| ≥15.2 kPa | 224 (12.6) | 91 (8.9) | 133 (17.6) |  | 0 | 224 (100) |  |
| **Any MRF, n (%)** | 1,225 (36.0) | 651 (34.6) | 574 (37.9) | *0.04* | 891 (38.5) | 194 (45.8) | *0.005* |
| **Cirrhosis, n (%)** | 424/2,736 (15.5) | 197/1,522 (12.9) | 227/1,214 (18.7) | *<0.001* | 2,312 (100) | 424 (100) | *na* |
| ** Cirrhosis=* defined as liver stiffness measurement ≥15.2 or platelets counts <150*10^9^ cells/L. Continuous variables are presented as median (IQR) unless stated otherwise.  ***Abbreviations****: sd: standard deviation; BMI= body mass index; ALT=alanine aminotransferase; AST= aspartate aminotransferase; GGT=gamma glutamyl transferase; ULN for ALT= upper limit of normal 41 IU/L for men and 31 IU/L for women; ULN for GGT=* ≥55 IU/L for men, ≥38 IU/L for women; *IQR: interquartile range; LSM=liver stiffness measurements; MRF=metabolic risk factor; na=not applicable.* | | | | | | | |

| **sTable 3: Baseline characteristics of all patients with CHD and CHB at initial HBsAg test** | | | | | |
| --- | --- | --- | --- | --- | --- |
| **Parameters** | **Number** | **All** | **CHD** | **CHB** | **p-value** |
| **Number** |  | 5,955 | 3,399 (57.1) | 2,556 (42.9) | <0.001 |
| **Sex, men, n (%)** | 5,955 | 2,857 (48.0) | 1,515 (44.6) | 1,342 (52.5) | <0.001 |
| Age at first HBsAg test, years, mean (sd) | 5,955 | 40.1 (10.7) | 42.7 (10.5) | 36.6 (9.9) | <0.001 |
| **Age, categories n (%)** | 5,955 |  |  |  |  |
| 18-29 |  | 1,000 (16.8) | 340 (10.0) | 660 (25.8) | <0.001 |
| 30-44 |  | 3,155 (53.0) | 1,726 (50.8) | 1,429 (55.9) | <0.001 |
| 45-59 |  | 1,518 (25.5) | 1,129 (33.2) | 389 (15.2) | 0.007 |
| ≥60 |  | 282 (4.7) | 204 (6.0) | 78 (3.1) | 0.23 |
| BMI, mean (sd) | 2,029 | 26.7 (4.7) | 26.8 (4.7) | 26.6 (4.6) | 0.25 |
| **Laboratory parameters** |  |  |  |  |  |
| ALT, IU/L, median (IQR) | 3,935 | 44.5 (27.4-78.1) | 56.6 (36.5-92.1) | 29.5 (19.6-48.0) | <0.001 |
| **ALT level categories, n (%)** | 3,935 | 3,935 | *2,382* | 1,553 |  |
| Normal ALT level, <1ULN |  | 1,478 (37.6) | 511 (21.5) | 967 (62.3) | <0.001 |
| ≥*1ULN-<2*ULN |  | 1,358 (34.5) | 983 (41.3) | 375 (24.1) | 0.06 |
| ≥2*ULN-<5*ULN |  | 891 (22.6) | 721 (30.3) | 170 (10.9) | 0.002 |
| ≥5*ULN-<10*ULN |  | 170 (4.3) | 141 (5.9) | 29 (1.9) | 0.02 |
| ≥10*ULN |  | 38 (1.0) | 26 (1.1) | 12 (0.8) | 0.08 |
| AST, IU/L, median (IQR) | 3,929 | 33.6 (22.8-52.8) | 42.1 (30.4-63.9) | 22.7 (18.1-32.0) | <0.001 |
| Albumin, g/L, median (IQR) | 3,411 | 42.8 (40.3-45.0) | 42.0 (39.6-44.1) | 43.9 (41.9-46.0) | 0.48 |
| Total bilirubin, µmol/L, median (IQR) | 3,639 | 12.6 (9.3-16.5) | 12.9 (9.6-16.8) | 12.0 (9.0-15.9) | 0.02 |
| Platelets count, 10*9 cells/L, median (IQR) | 3,343 | 214.7 (176.0-257.0) | 198.7 (159.0-236.0) | 244.0 (208.0-282.0) | <0.001 |
| Platelets count <150 10*9 cells/L, n (%) | 3,343 | 448 (13.4) | 416/2,092 (19.9) | 40/1,251 (3.2) | <0.001 |
| GGT, IU/L, median (IQR) | 3,019 | 34.5 (21.8-57.8) | 39.6 (25.4-65.0) | 26.7 (18.1-46.9) | <0.001 |
| Elevated GGT,≥*1ULN, n (%) | 3,019 | 1,046 (34.6) | 779/1,850 (42.1) | 267/1,169 (22.8) | <0.001 |
| **Virological parameters** |  |  |  |  |  |
| HBeAg, positive, n (%) | 1,437 | 519 (36.1) | 306/810 (37.8) | 213/627 (34.0) | 0.14 |
| HBsAg log_10_ IU/mL, median (IQR) | 3,571 | 3.7 (3.1-4.0) | 3.8 (3.4-4.1) | 3.4 (2.7-3.8) | <0.001 |
| HBV DNA log_10_ IU/mL, median (IQR) | 3,821 | 5.4 (4.3-6.2) | 2.2 (1.3-3.2) | 3.3 (2.4-4.2) | <0.001 |
| HDV RNA log_10_ IU/mL, median (IQR) | 2,463 | 2.7 (1.7-3.7) | 5.4 (4.3-6.2) | na | na |
| LSM, kPa, median (IQR) | 2,446 | 7.2 (5.3-10.2) | 8.2 (6.1-11.4) | 5.1 (4.2-6.8) | <0.001 |
| **LSM, categories, n (%)** |  | 2,446 | 1,782 | 664 |  |
| F0-F1 |  | 1,277 (52.2) | 740 (41.5) | 537 (80.94) | <0.001 |
| F2 |  | 522 (21.3) | 475 (26.7) | 47 (7.1) | <0.001 |
| F3 |  | 289 (15.9) | 343 (19.2) | 46 (6.9) | <0.001 |
| F4 |  | 258 (10.5) | 224 (12.6) | 34 (5.1) | 0.40 |
| **Any MRF, n (%)** | 5,955 | 1,929 (32.4) | 1,225 (36.0) | 704 (27.5) | <0.001 |
| **Cirrhosis*, n (%)** | 4,267 | 483 (11.3) | 424/2,736 (15.5) | 59/1,531 (3.9) | <0.001 |
| *Cirrhosis= defined as liver stiffness measurement ≥15.2 or platelets counts <150*10^9^ cells/L.  ***Abbreviations****: sd: standard deviation; IQR=25^th^ – 75^th^ interquartiles; BMI= body mass index; ALT=alanine aminotransferase; AST= aspartate aminotransferase; GGT=gamma glutamyl transferase; ULN= upper limit of normal 41 IU/L for men and 31 IU/L for women; LSM=liver stiffness measurements; MRF=metabolic risk factor; na=not applicable.* | | | | | |

| **sTable 4: Parameters associated with being diagnosed with CHD versus CHB in univariable regression models (all study participants). Crude Odds ratio (OR) and (95% confidence intervals, CI) are presented.** | | | | | |
| --- | --- | --- | --- | --- | --- |
| **Age groups** | **All** | **18-29** | **30-44** | **45-59** | **≥60** |
| **Variables** |  | **Crude OR (95% CI)** | | | |
| Sex (Women vs men) | 1.38 (1.24-1.52) | 1.10 (0.85-1.43) | 1.05 (0.91-1.20) | 1.75 (1.39-2.21) | 2.38 (1.37-4.15) |
| BMI>23.0 | 0.88 (0.71-1.09) | 0.70 (0.42-1.12) | 1.18 (0.87-1.60) | 1.99 (0.99-3.99) | 1.58 (0.48-5.27) |
| **Laboratory parameters** |  |  |  |  |  |
| ALT above ULN | 6.04 (5.24-6.96) | 8.04 (5.29-12.22) | 6.20 (5.12-7.51) | 5.51 (4.03-7.53) | 5.74 (2.78-11.85) |
| AST above ULN | 7.90 (6.82-9.16) | 10.56 (7.16-15.57) | 7.53 (6.16-9.21) | 6.37 (4.63-8.77) | 5.28 (2.56-10.86) |
| Albumin level <30 g/L | 3.78 (1.78-8.03) | 2.78 (0.46-16.79) | 4.66 (1.38-15.73) | na | 0.33 (0.07-1.72) |
| Total bilirubin >21 µmol/L | 1.30 (1.04-1.62) | 0.91 (0.51-1.63) | 1.31 (0.98-1.75) | 1.57 (0.90-2.75) | 2.33 (0.65-8.31) |
| Platelet count <150 x 10*9 cells/L | 7.52 (5.39-10.48) | 5.70 (2.25-14.46) | 9.55 (5.59-16.32) | 5.24 (2.92-9.42) | 1.44 (0.54-3.84) |
| Elevated GGT, ≥55 IU/L in men,≥38 IU/L in women | 2.46 (2.08-2.90) | 2.89 (1.83-4.56) | 2.02 (1.63-2.52) | 2.40 (1.66-3.48) | 1.33 (0.62-2.86) |
| **Virological parameters** |  |  | | | |
| HBeAg, positive | 1.18 (1.00-1.47) | 0.81 (0.52-1.27) | 1.64 (1.21-2.22) | 2.12 (1.03-4.34) | 1.43 (0.44-4.66) |
| HBsAg ≥10,000 IU/mL | 2.29 (1.94-2.70) | 1.60 (1.09-2.35) | 2.82 (2.26-3.51) | 4.31 (2.55-7.27) | 6.00 (1.37-26.36) |
| HBV DNA <2,000 IU/mL | 3.41 (2.96-3.92) | 3.40 (2.27-5.10) | 3.58 (2.95-4.34) | 3.70 ( 2.72-5.02) | 1.89 (0.94-3.80) |
| **LSM, categories** |  |  | | | |
| LSM <7.5 kPa | 0.17 (0.14-0.21) | 0.27 (0.16-0.46) | 0.13 (0.10-0.18) | 0.30 (0.19-0.45) | 0.29 (0.10-0.78) |
| LSM ≥12.5 kPa | 4.51 (3.14-6.50) | 3.01 (1.11-8.49) | 5.43 (3.15-9.35) | 2.27 (1.24-4.18) | 8.83 (1.13-68.79) |
| **Any MRF** | 1.48 (1.33-1.66) | 1.60 (1.16-2.21) | 1.18 (1.01-1.37) | 1.43 (1.12-1.82) | 1.02 (0.59-1.75) |
| **Cirrhosis*** | 4.58 (3.46-6.05) | 4.15 (1.73-9.97) | 5.33 (3.45-8.23) | 2.54 (1.61-4.02) | 2.20 (0.81-6.01) |
| * Cirrhosis= defined as liver stiffness measurement ≥15.2 for CHD and ≥12.5 for CHB or platelets counts <150*10^9^ cells/L.  Abbreviations: CHD=chronic hepatitis D; BMI= body mass index; ALT=alanine aminotransferase; AST= aspartate aminotransferase; GGT=gamma glutamyl transferase; ULN= upper limit of normal; LSM=liver stiffness measurements; MRF=metabolic risk factor | | | | | |

| **sTable 5: Parameters associated with LSM ≥15.2 kPa in patients with CHD (n=3,399) in univariable and multivariable logistic regression models. Crude Odds ratio (OR) and adjusted OR (95% confidence intervals, CI) are presented.** | | | | |
| --- | --- | --- | --- | --- |
| **Parameters** | **OR (95% CI)** | **p-value** | **aOR (95% CI)** | **p-value** |
| Sex, men vs women | 2.19 (1.65-2.92) | <0.001 | 1.97 (1.11-3.50) | 0.02 |
| Age <30 years of age | 0.40 (0.21-0.76) | 0.006 | 0.97 (0.92-1.02) | 0.27 |
| BMI ≥23.0 | 5.63 (2.43-13.03) | <0.001 | 3.78 (0.77-18.57) | 0.10 |
| Laboratory parameters |  |  |  |  |
| ALT level, ≥1*ULN | 4.42 (2.36-8.28) | <0.001 | 1.61 (0.56-4.62) | 0.37 |
| AST level ≥1*ULN | 5.30 (3.08-9.15) | <0.001 | 2.56 (1.00-6.60) | 0.05 |
| Low albumin level than 30 g/L | 6.22 (2.60-14.90) | <0.001 | 4.13 (0.99-17.28) | 0.05 |
| Total bilirubin >21 µmol/L | 4.03 (2.64-6.17) | <0.001 | 2.94 (1.52-5.70) | <0.001 |
| Elevated GGT ≥1*ULN | 6.92 (4.49-10.65) | <0.001 | 5.29 (2.83-9.87) | 0.001 |
| Platelets count <150 10*9 cells/L | 7.48 (5.19-10.78) | <0.001 | 5.73 (3.24-10.11) | <0.001 |
| Virological parameters |  |  |  |  |
| HbeAg, positive | 1.04 (0.53-2.03) | 0.91 |  |  |
| HBsAg ≥10,000 IU/mL | 0.55 (0.37-0.83) | 0.004 | 0.71 (0.40-1.23) | 0.21 |
| HDV RNA ≥100,000 IU/mL | 1.68 (1.16-2.43) | 0.006 | 2.38 (1.32-4.27) | 0.004 |
| HBV DNA <2,000 IU/mL | 1.18 (0.73-1.90) | 0.49 |  |  |
| Any MRF | 1.75 (1.32-2.32) | <0.001 | 2.00 (1.20-3.30) | 0.008 |
| **Abbreviations**: BMI= body mass index; ALT=alanine aminotransferase; AST= aspartate aminotransferase; GGT=gamma glutamyl transferase; ULN= upper limit of normal; In men the ULN for ALT was 41 IU/L, was 35 IU/L for AST, was 55 IU/L for GGT and in women 31 IU/L, 31 IU/L and 38 IU/L respectively; MRF=metabolic risk factor; Multivariable model adjusted for sex, age, BMI, ALT, AST, albumin, bilirubin, GGT, platelets, HBsAg, HDV RNA, and any MRF. | | | | |

| **sTable 6: Parameters associated with HDV RNA ≥100,000 IU/mL in patients with CHD (n=3,399) in univariable and multivariable regression models. Crude Odds ratio (OR) and adjusted OR (95% confidence intervals, CI) are presented.** | | | | |
| --- | --- | --- | --- | --- |
| **Variables** | **Crude OR (95% CI)** | **p-value** | **aOR (95% CI)** | **p-value** |
| Sex, men | 1.22 (1.04-1.43) | 0.02 | 0.70 (0.43-1.10) | 0.12 |
| Age <30 years of age | 1.19 (0.89-1.59) | 0.25 |  |  |
| BMI categories |  |  |  |  |
| Overweight | 1.04 (0.75-1.45) | 0.80 |  |  |
| Obesity | 1.05 (0.80-1.37) | 0.74 |  |  |
| Laboratory parameters, |  |  |  |  |
| ALT level, ≥1*ULN | 2.71 (2.17-3.37) | <0.001 | 0.83 (0.40-1.72) | 0.61 |
| AST level, ≥1*ULN | 1.92 (1.58-2.33) | <0.001 | 1.65 (0.89-3.07) | 0.11 |
| Albumin level <30 g/L | 1.79 (0.91-3.53) | 0.09 |  |  |
| Total bilirubin >21 µmol/L | 1.54 (1.17-2.02) | 0.53 |  |  |
| Platelet count <150 10*9 cells/L | 1.27 (1.00-1.61) | 0.05 |  |  |
| Elevated GGT, ≥55 IU/L in men, ≥38 IU/L in women | 1.34 (1.09-1.64) | 0.005 | 1.35 (0.80-2.30) | 0.26 |
| Virological parameters |  |  |  |  |
| HBeAg, positive | 1.56 (1.09-2.23) | 0.02 | 1.40 (0.87-2.27) | 0.17 |
| HBsAg ≥10,000 IU/mL | 2.58 (2.10-3.19) | <0.001 | 2.79 (1.68-4.64) | <0.001 |
| HBV DNA <2,000 IU/mL | 0.95 (0.75-1.21) | 0.68 |  |  |
| LSM ≥15.2 kPa | 1.68 (1.16-2.43) | 0.006 |  |  |
| Any MRF | 0.96 (0.82-1.14) | 0.65 |  |  |
| Cirrhosis* | 1.41 (1.10-1.81) | 0.006 | 1.17 (0.54-2.53) | 0.70 |
| *Cirrhosis= defined as liver stiffness measurement ≥15.2 or platelets counts <150*10^9^ cells/L. Abbreviations: CHD=chronic hepatitis D; sd: standard deviation; BMI= body mass index; ALT=alanine aminotransferase; AST= aspartate aminotransferase; GGT=gamma glutamyl transferase; ULN= upper limit of normal; LSM=liver stiffness measurements; MRF= metabolic risk factor. Multivariable model adjusted for sex, ALT, AST, GGT, HBeAg, HBsAg, and cirrhosis. | | | | |

| **sTable 7: Parameters associated with HBsAg ≥10,000 IU/mL in patients with CHD (n=3,399) in univariable and multivariable regression models. Crude Odds ratio (OR) and adjusted OR (95% confidence intervals, CI) are presented.** | | | | |
| --- | --- | --- | --- | --- |
| **Variables** | **Crude OR (95% CI)** | **p-value** | **aOR (95% CI)** | **p-value** |
| Sex, women vs men | 1.28 (1.07-1.53) | 0.008 | 1.64 (0.97-2.78) | 0.06 |
| Age <30 years of age | 0.75 (0.55-1.03) | 0.08 |  |  |
| BMI ≥23.0 | 1.12 (0.84-1.50) | 0.45 |  |  |
| Laboratory parameters | | | | |
| ALT level, ≥1*ULN | 1.29 (1.01-1.63) | 0.04 | 1.56 (0.77-3.17) | 0.21 |
| AST level, ≥1*ULN | 1.00 (0.82-1.23) | 0.97 |  |  |
| Albumin level <30 g/L | 0.63 (0.28-1.40) | 0.25 |  |  |
| Total bilirubin >21 µmol/L | 0.57 (0.41-0.80) | <0.001 | 0.40 (0.15-1.06) | 0.07 |
| Platelet count <150 10*9 cells/L | 0.76 (0.59-0.98) | 0.03 |  |  |
| Elevated GGT, ≥55 IU/L in men, ≥38 IU/L in women | 0.70 (0.57-0.87) | 0.001 | 0.48 (0.27-0.88) | 0.02 |
| Virological parameters | | | | |
| HBeAg, positive | 2.56 (1.80-3.62) | <0.001 | 1.98 (1.15-3.42) | 0.01 |
| HDV RNA ≥100,000 IU/mL | 2.58 (2.10-3.19) | <0.001 | 2.79 (1.59-4.91) | <0.001 |
| HBV DNA <2,000 IU/ml | 0.86 (0.67-1.11) | 0.24 |  |  |
| LSM categories |  |  |  |  |
| LSM <7.5 kPa | 1.14 (0.89-1.46) | 0.29 |  |  |
| LSM ≥15.2 kPa | 0.55 (0.37-0.83) | 0.004 |  |  |
| Any MRF | 0.97 (0.81-1.17) | 0.74 |  |  |
| Cirrhosis* | 0.68 (0.52-0.89) | 0.005 | 0.81 (0.32-2.07) | 0.66 |
| *Cirrhosis= defined as liver stiffness measurement ≥15.2 or platelets counts <150*10^9^ cells/L. Abbreviations: CHD=chronic hepatitis D; sd: standard deviation; BMI= body mass index; ALT=alanine aminotransferase; AST= aspartate aminotransferase; GGT=gamma glutamyl transferase; ULN= upper limit of normal; IQR: interquartile range; LSM=liver stiffness measurements; MRF=metabolic risk factor. Multivariable model adjusted for sex, bilirubin, GGT, HBeAg, HDV RNA, and cirrhosis. | | | | |

| **sTable 8: Baseline characteristics of 2,231 pairs of individuals with CHD and CHB matched through propensity scores on sex and age at HBsAg test** | | | | | |
| --- | --- | --- | --- | --- | --- |
|  | **Variables** | **Number** | **CHD** | **CHB** | **p-value** |
|  | **Number of patients** |  | 2,231 (100) | 2,231 (100) |  |
|  | **Sex, men** | 2,231 | 1,137 (51.0) | 1,137 (51.0) | 1.0 |
|  | **Age** | 2,231 |  |  |  |
|  | Age at first HBsAg test all, years, mean (sd) |  | 38.3 (9.3) | 37.8 (9.6) | 0.06 |
|  | Age, men, years, mean (sd) |  | 37.7 (8.6) | 37.1 (9.0) | 0.11 |
|  | Age, men with cirrhosis, years, mean (sd) |  | 39.4 (8.3) | 41.9 (10.1) | <0.001 |
|  | Age, women, years mean (sd) |  | 39.0 (9.8) | 38.5 (10.2) | 0.26 |
|  | Age, women with cirrhosis, years, mean (sd) |  | 42.2 (47.6) | 47.6 (12.0) | <0.001 |
|  | **Age, categories, n (%)** | 2,231 |  |  |  |
|  | 18-29 |  | 340 (15.2) | 443 (19.9) | <0.001 |
|  | 30-44 |  | 1,430 (64.1) | 1,327 (59.5) | 0.002 |
|  | 45-59 |  | 389 (17.4) | 389 (17.4) | 1.0 |
|  | ≥60 |  | 72 (3.2) | 72 (3.2) | 1.0 |
|  | BMI, mean (sd) |  | 26.4 (4.7) | 26.7 (4.6) | 0.13 |
|  | **BMI categories, n (%)** | 1,458 | 806 | 652 |  |
|  | Underweight |  | 14 (1.7) | 11 (1.7) | 0.94 |
|  | Normal |  | 190 (23.6) | 128 (19.6) | 0.07 |
|  | Overweight |  | 301 (37.3) | 250 (38.3) | 0.70 |
|  | Obesity |  | 301 (37.3) | 263 (41.9) | 0.24 |
|  | **Laboratory parameters** |  |  |  |  |
|  | ALT, IU/L, median (IQR) | 2,907 | 58.5 (37.8-95.2) | 29.0 (19.4-47.6) | <0.001 |
|  | **ALT level categories adjusted** | 2,907 | 1,546 | 1,361 |  |
|  | Normal ALT level, <1ULN |  | 336 (21.7) | 852 (62.6) | <0.001 |
|  | ≥*1ULN-<2*ULN |  | 132 (40.4) | 329 (24.2) | 0.001 |
|  | ≥2*ULN-<5*ULN |  | 472 (30.5) | 146 (10.7) | <0.001 |
|  | ≥5*ULN-<10*ULN |  | 90 (5.8) | 27 (2.0) | <0.001 |
|  | ≥10*ULN |  | 22 (1.4) | 7 (0.5) | 0.01 |
|  | AST, IU/L, median (IQR) | 2,903 | 41.7 (30.1-63.2) | 22.7 (18.0-31.7) | <0.001 |
|  | Albumin, g/L, median (IQR) | 2,514 | 42.3 (40.0-44.5) | 43.9 (41.9-46.0) | 0.38 |
|  | Total bilirubin, µmol/L, median (IQR) | 2,690 | 12.9 (9.5-16.8) | 11.9 (9.0-15.7) | 0.04 |
|  | Platelets count, 10*9 cells/L, median (IQR) | 2,479 | 204.0 (167.1-240.4) | 244.0 (207.0-281.0) | <0.001 |
|  | Platelets count <150 10*9 cells/L | 2,479 | 228/1,368 (16.7) | 37/1,111 (3.3) | <0.001 |
|  | GGT, IU/L, median (IQR) | 2,231 | 38.9 (25.0-63.2) | 26.6 (18.0-47.6) | 0.001 |
|  | Elevated GGT ≥*1ULN | 2,231 | 477/1,208 (39.5) | 240/1,023 (23.5) | <0.001 |
|  | **Virological parameters** |  |  |  |  |
|  | HbeAg, positive, n (%) | 1,092 | 224/571 (39.2) | 161/521 (30.9) | 0.004 |
|  | HBsAg log_10_ IU/mL, median (IQR) | 2,630 | 3.9 (3.4-4.1) | 3.3 (2.6-3.8) | <0.001 |
|  | HBV DNA log_10_ IU/mL, median (IQR) | 2,860 | 2.3 (1.4-3.2) | 3.3 (2.5-4.2) | <0.001 |
|  | HDV RNA log_10_ IU/mL, median (IQR) | 1,592 | 5.4 (4.3-6.2) | na | na |
|  | LSM, kPa, median (IQR) | 1,762 | 8.0 (6.1-10.9) | 5.1 (4.2-6.8) | <0.001 |
|  | **LSM, categories, n (%)** | 1,762 | 1,166 | 596 |  |
|  | F0-F1 |  | 514 (44.1) | 479 (80.4) | <0.001 |
|  | F2 |  | 304 (26.1) | 42 (7.0) | <0.001 |
|  | F3 |  | 214 (18.4) | 44 (7.4) | <0.001 |
|  | F4 |  | 134 (11.5) | 31 (5.2) | <0.001 |
|  | Any MRF, n (%) | 4,462 | 719 (32.2) | 638 (28.6) | 0.008 |
|  | Cirrhosis, n (%) | 3,145 | 237/1,790 (13.2) | 54/1,355 (4.0) | <0.001 |
|  | * Cirrhosis= defined as liver stiffness measurement ≥15.2 or platelets counts <150*10^9^ cells/L.  **Abbreviations**: sd= standard deviation; IQR= 25^th^ – 75^th^ interquartile range; BMI= body mass index; ALT=alanine aminotransferase; AST= aspartate aminotransferase; GGT=gamma glutamyl transferase; ULN= upper limit of normal 41 IU/L for men and 31 IU/L for women; LSM=liver stiffness measurements; F0-F4= fibrosis stage see methods for cutoffs; MRF=metabolic risk factor; na=not applicable. | | | | |

| **sTable 9: Parameters associated with diagnosis of CHD in all propensity scores matched individuals (n=4,462) in univariable and multivariable logistic regression models. Crude Odds ratio (OR) and adjusted OR (95% confidence intervals, CI) are presented.** | | | | |
| --- | --- | --- | --- | --- |
| **Variables** | **OR (95% CI)** | **p-value** | **aOR (95% CI)** | **p-value** |
| BMI ≥23.0 | 0.81 (0.63-1.03) | 0.09 |  |  |
| Laboratory parameters | | | | |
| ALT level, ≥1*ULN | 3.49 (3.02-4.02) | <0.001 | 2.58 (1.32-5.06) | 0.006 |
| AST level ≥1*ULN | 7.51 (6.36-8.87) | <0.001 | 5.49 (2.92-10.30) | <0.001 |
| Albumin level <30 g/L | 3.60 (1.57-8.25) | <0.001 |  |  |
| Total bilirubin >21 µmol/L | 1.30 (1.04-1.62) | 0.02 |  |  |
| Elevated GGT ≥1*ULN | 2.13 (1.77-2.56) | <0.001 | 0.54 (0.29-1.00) | 0.05 |
| Platelets count <150 10*9 cells/L | 5.81 (4.06-8.30) | <0.001 | 4.27 (1.25-14.56) | 0.02 |
| Virological parameters | | | | |
| HBeAg, positive | 1.44 (1.12-1.85) | 0.004 | 2.46 (1.33-4.55) | 0.004 |
| HBsAg ≥10,000 IU/mL | 2.72 (2.25-3.28) | <0.001 | 1.18 (0.68-2.04) | 0.55 |
| HBV DNA <2,000 IU/mL | 3.40 (2.89-4.01) | <0.001 | 6.18 (3.39-11.26) | <0.001 |
| LSM categories | | | | |
| LSM ≥7.5 kPa | 5.19 (4.11-6.56) | <0.001 |  |  |
| LSM ≥12.5 kPa | 4.07 (2.76-6.02) | <0.001 |  |  |
| Any MRF | 1.19 (1.05-1.35) | 0.008 | 1.13 (0.70-1.84) | 0.62 |
| Cirrhosis* | 3.68 (2.71-4.99) | <0.001 | 1.36 (0.75-2.47) | 0.31** |
| *Cirrhosis= defined as liver stiffness measurement ≥15.2 for CHD and ≥12.5 for CHB or platelets counts <150*10^9^ cells/L. **Tested when platelets not included in the model.  Abbreviations: CHD=chronic hepatitis D; BMI= body mass index; ALT=alanine aminotransferase; AST= aspartate aminotransferase; GGT=gamma glutamyl transferase; ULN= upper limit of normal; in men the ULN for ALT was 41 IU/L, was 35 IU/L for AST, was 55 IU/L for GGT and in women 31 IU/L, 31 IU/L and 38 IU/L respectively; LSM=liver stiffness measurements; MRF=metabolic risk factor, Multivariable model adjusted for ALT, AST, albumin, bilirubin, GGT, HBeAg, HBsAg, HBV DNA, and MRF. | | | | |

**References (supplements):**
